# Ultra-processed food exposure and cognitive outcomes: A systematic review of observational studies

**DOI:** 10.1101/2025.02.12.25322127

**Authors:** Megan Smith, Pippa Watson, John Gallacher, Sarah Bauermeister

## Abstract

**Background:** Ultra-processed food (UPF) intake has been associated with multiple negative health outcomes. Research investigating UPF intake and cognitive health outcomes has begun. The aim of this review is to summarise the existing evidence of associations between exposure to UPFs, as defined by the NOVA food classification system, and cognitive health outcomes.

**Methods:** We conducted a systematic search across MEDLINE, PsycINFO, Embase, and APA Psych Articles through OVID, and PubMed for relevant studies up-until October 2024. The Newcastle-Ottawa Scale was used to assess the quality of included studies. A narrative approach was used to summarise and integrate results across studies.

**Results:** Three hundred and eighty-three articles were screened and five met the inclusion criteria. All studies were published between 2022 and 2024. The association between UPF intake and four different cognitive outcomes (dementia risk, cognitive impairment risk, cognitive performance and cognitive change trajectories) were explored across the studies. Three out of the five included studies found a significant negative main effect of consuming UPF on the cognitive outcome of interest. All studies identified adverse consequences of consumption in either a sub-group of the population or a sub-group of UPF type.

**Conclusions:** Deleterious effects of UPF consumption on multiple cognitive health outcomes were identified across all studies. However, the results suggest the relationship may be specific to sub-groups of the population or sub-groups of UPF type. Conclusions should be drawn with caution due to the limited number of studies available examining UPF intake according to NOVA and its association with cognitive outcomes, as well as the variability in cognitive measures assessed and other methodological differences across studies.

## Introduction

Currently, more than 55 million people are living with dementia world-wide (GBD Dementia Forecasting Collaborators., 2022). The social, economic and healthcare burden of dementia is vast, due to those living with the condition often experiencing a reduction in independence (World Health Organisation, 2023). Prevention strategies and risk reduction play an integral role in managing the incidence of dementia and cognitive decline. These strategies assist by slowing or averting the neurodegenerative processes that lead to cognitive decline and dementia. Many risk factors are lifestyle or “modifiable” factors, such as diabetes, cholesterol and hypertension (Livingston et al., 2024). Several of these modifiable risk factors can be adjusted by diet, meaning a person’s risk of developing cognitive decline or dementia may be increased, or decreased, through factors related to what they eat and drink. For instance, an unhealthy diet can contribute to the development of type 2 diabetes (a risk factor for dementia) whilst a balanced, nutrient-dense healthy diet can prevent diabetes onset (Hu., 2011) and thus mitigate the associated dementia risk.

Whilst multiple definitions of ultra-processed foods (UPFs) exist, the most widely used is that defined by the NOVA classification system (Monterio et al., 2018). The NOVA system is a framework which classifies all food items according to its degree and purpose of processing. According to this classification, UPFs are manufactured items containing minimal whole foods, alongside numerous substances that are of little or no culinary use (e.g. high-fructose corn syrup and modified starches) which are added to enhance the flavour, texture, and/or the shelf-life of the product. Ultra-processed foods are now the main – or a major – source of dietary energy in many high-income countries including the United Kingdom (UK) and United States of America (Rauber et al., 2021; Zhang et al., 2021). Consumption patterns in low to middle-income countries are more recently demonstrating the shift away from traditional diets and towards UPFs (Baker et al., 2020).

Large-scale epidemiological studies highlight an increased risk of nearly all major non-communicable diseases are associated with high intakes of UPFs. For instance, high UPF consumption has been associated with an increased risk of all-cause mortality (Rico-Campà et al., 2019), cardiovascular disorders (Srour et al., 2019), type 2 diabetes (Dicken et al., 2024), cancers (Chang et al., 2023) and obesity (Rauber et al., 2021) risk. A recent umbrella review found greater exposure to UPFs was associated with a higher risk of adverse health outcomes (Lane et al., 2024). Discussions regarding public health strategies and policies to limit UPF intake have consequently become salient. Systematic reviews aid the debate by consolidating evidence, identifying knowledge gaps and clarifying conflicting findings.

It is plausible that UPF intake might influence cognitive health and dementia risk as biological pathways implicated in maintaining brain health, such as inflammation and the gut microbiome, are modulated by UPF intake (Silva dos Santos et al., 2023; Cuevas-Sierra et al., 2021). However, investigations on the cognitive effects of consuming UPFs has been initiated with conflicting results.

So far, two systematic reviews exist summarising the evidence regarding UPF intake and dementia (Henney et al., 2024) and Alzheimer’s Disease (Claudino et al., 2024) risk. Further numerous reviews have assessed the associations between individual UPF items and cognitive health outcomes, for instance sugar-sweetened beverages and cognitive disorders (Sun et al., 2022). However, no review has accumulated the evidence regarding total UPF intake and cognitive health outcomes. Therefore, we aim to bridge this gap and collate, summarise and evaluate all existing evidence regarding the exposure of UPFs and cognitive functioning, cognitive decline, and dementia risk. Evidence regarding this topic has amassed lately as the debate of UPFs contribution to adverse health outcomes has gained traction both publicly and scientifically in recent years; a review is timely. Uniquely, this report will only consider research that has classified the degree of food processing using the NOVA classification system. By doing so, this review may provide valuable insights which could inform development of public policies and strategies as the UK government has stated the NOVA framework is the only current classification system that is applicable in the UK.

## Methods

We conducted and reported this systematic review in accordance with the Preferred Reporting Items for Systematic Reviews and Meta-Analyses (PRISMA) guidelines. The systematic review was pre-registered on PROSPERO (CRD42024600338).

### Inclusion criteria

The inclusion criteria were constructed in line with the population, exposure, comparisons, outcomes and study design (PECOS) reporting structure. Therefore, studies involving human adults (over 18 years of age) who were free from dementia at baseline (population) were eligible. The intake of UPFs needed to be measured and defined by the NOVA classification system (exposure). The comparison required was higher versus lower UPF exposure. Studies were required to investigate associations between intake and cognitive performance, cognitive performance trajectories, or dementia risk (outcome).

Epidemiological observational studies, including prospective cohort, case-control, and cross-sectional studies (study design), were included. The full exclusion and inclusion criteria are in supplementary table 1.

Studies which measured the intake of only one type of UPF (e.g. sugar sweetened beverages), or measured a diet that is high in UPF but did not specifically investigate overall UPF intake (e.g. studies which explored “western diet patterns”, which are characteristically high in UPFs but include other non-UPF items), were not included in this review.

### Search strategy

The lead author conducted a systematic search across MEDLINE, PsycINFO, Embase, and APA Psych Articles through OVID, and PubMed for relevant studies up-until October 2024. Additionally, the reference list of included articles was scanned to identify potentially relevant studies not captured by the search. The search terms were words relevant to ultra-processed foods, NOVA classification and cognitive outcomes (full search terms in supplementary table 2).

The identified studies were exported to Rayyan software. Duplicates were removed first before two reviewers independently determined eligible studies. The initial selection was conducted based on titles and abstracts, with studies deemed potentially relevant being read in full before being included or excluded. Any disagreements between reviewers were resolved by a third reviewer.

### Data extraction

One reviewer extracted data from the studies included using a pre-determined custom Microsoft Excel Spreadsheet. Extracted data included study characteristics such as year of publication, country, sample size, sample characteristics (e.g. % female, mean age). Data relevant to the methods, such as dietary intake assessment, dementia diagnosis or cognitive performance measurement, and statistical methods utilised was extracted. The results obtained between assessing UPF intake and dementia risk or cognitive performance (i.e. hazards or odds ratio and 95% confidence intervals, or beta-values for association scores and 95% confidence intervals), were extracted. Additionally, factors required to evaluate the quality of the studies were obtained (e.g. follow up duration and adjustment of confounders).

### Quality of evidence

To assess the quality of evidence in the studies included we applied the Newcastle–Ottawa scale (NOS; Wells et al., 2015). This tool consists of eight items, evaluating the quality of the study over three domains (selection, comparability and outcome). The total maximum score is nine (one point available for all but one item, where two points are available). Studies obtaining scores lower than five points were classified as poor-quality, those with five or six points as medium quality and those with seven or more points were considered high quality (Delpino et al., 2022; Henney et al., 2024). Where appropriate, we utilised the adapted version of the NOS that has been developed for the specific assessment of cross-cohort studies. Two reviewers independently rated the included evidence, with any disagreement resolved by third reviewer.

### Data synthesis

To integrate the findings of the included studies we used a narrative approach. This method was chosen due to the small number of studies and the heterogeneity in outcomes investigated. Synthesis was initially assessed across all studies to examine the overarching theme of UPF and cognitive outcomes (e.g., dementia risk, cognitive performance and cognitive change). Studies were then grouped according to the cognitive outcome of interest to evaluate the evidence pertinent to these outcomes.

## Results

### Study selection and characteristics

A flowchart demonstrating the selection process of the studies is illustrated in Figure 1. After removal of duplicates (n = 255), 383 articles were screened and five met the inclusion criteria.

**Figure 1.**
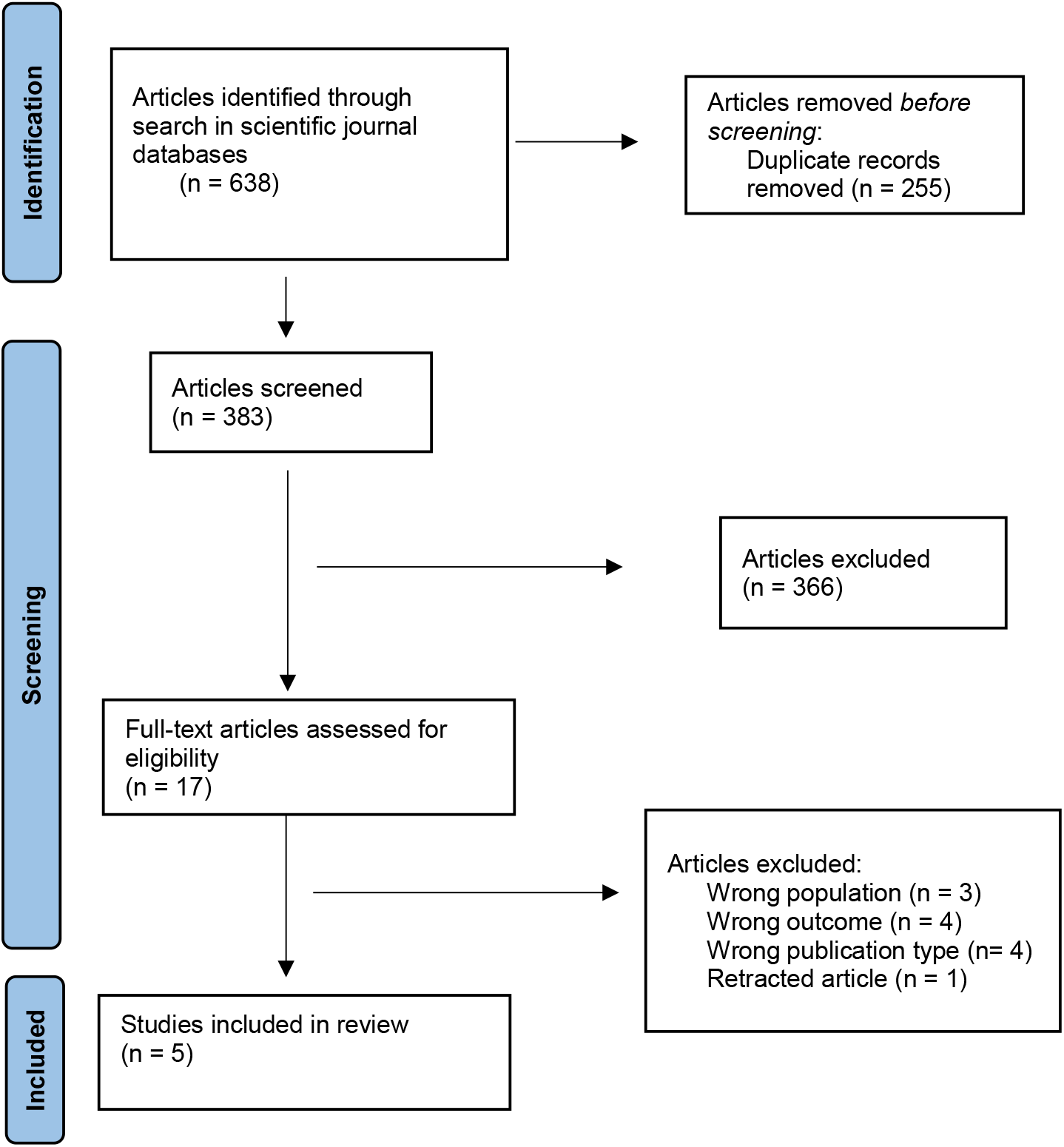
Preferred reporting items for systematic reviews and meta-analysis (PRISMA) flow diagram for study selection process.

The included studies (table 1) were conducted in America (Cardosa et al., 2022; Bhave et al., 2024), the UK (Li et al., 2022), Israel (Weinstein et al., 2023) and Brazil (Gonçalves et al., 2023). All studies were published recently to this review, between 2022 – 2024. One study was cross-sectional (Cordosa et al., 2022) and four (Bhave et al., 2024; Li et al., 2022; Weinstein et al., 2023; Gonçalves et al., 2023) were longitudinal, with follow-ups ranging from 5.3 (Weinstein et al., 2023) to 10 years (Li et al., 2022).

**Table 1:** Characteristics of Included studies.

| Authors, Year | Country (cohort) | Study design | Sample Size | Follow up duration Years | Age (mean, sd) | Sex (% female) | Dietary data collection method | Level of exposure (expressed as) |
| --- | --- | --- | --- | --- | --- | --- | --- | --- |
| Cardoso et al., 2022 | America (NHANES) | Cross-sectional cohort | 2713 | N/A | 69.1 (0.2) | 54% female | 24-hr dietary recall | Tertiles (% kcal/day) |
| Li et al., 2022 | UK (The UK Biobank) | Prospective cohort | 72,083 | Median 10 | 61.6 (3.94) | 52.9 | 24-hr dietary recall | Continuous 10% increase (% g/day) |
| Weinstien et al., 2023 | Israel (IDCD) | Prospective cohort | 568 | Mean 5.3 | 71.3 (4.5) | 40 | FFQ | Quartiles (% kcal/day) |
| Gonçalves et al., 2023 | Brazil (ELSA-Brazil) | Prospective cohort | 10,775 | Median 8 | 51.6 (8.9) | 54.6 | FFQ | Quartiles (% kcal/day) |
| Bhave et al., 2024 | America (REGARDS) | Prospective cohort | 14,175 | Mean 10.7 | 63.5 (8.6) | 58 | FFQ | Continuous 10% increase (% g/day) |

NHANES: National Health and Nutrition Examination Survey; IDCD: Israel Diabetes and Cognitive Decline; ELSA-Brazil: Brazilian Longitudinal Study of Adult Health; REGARDS: Reasons for Geographic and Racial Differences in Stroke.

Sample sizes ranged from 568 (Weinstein et al., 2023) to 72,083 (Li et al., 2022). Three studies collected dietary data via Food Frequency Questionnaires (FFQ; Weinstein et al., 2023; Bhave et al., 2024; Gonçalves et al., 2023) and two (Li et al., 2022; Cordosa et al., 2022) used 24-hour dietary recall methods. All studies included an exposure variable of the percentage of the person’s diet that is ultra-processed, in some studies (Li et al., 2022; Bhave et al., 2024) this was expressed as a percentage of total weight (% g/day) whilst in others (Carodosa et al., 2022; Weinstein et al., 2023; Gonçalves et al., 2023) it was expressed as a percentage of total calorie intake (% kcal/day). Ultra-processed food intake was modelled as a continuous exposure variable (Li et al., 2022; Bhave et al., 2024) and as a categorical variable (Cardosa et al., 2022; Gonçalves et al., 2023; Weinstein et al., 2023). Across included studies, the association between UPF intake and a range of outcomes were explored: one study assessed the association with dementia risk (Li et al., 2022) , one study evaluated the association with cognitive impairment risk (Bhave et al., 2024), one study explored the relationship with cognitive performance (Cardosa et al., 2022), and three studies estimated the association with cognitive change (Bhave et al., 2024; Gonçalves et al., 2023; Weinstein et al., 2023). Across those measuring cognitive performance (n = 4), all employed a different number of tests, ranging from three (Cardosa et al., 2022) to 14 (Weinstein et al., 2023). Seventeen unique cognitive tests were administered across the studies, of these, 10 tests were conducted in more than one study. All studies controlled for age, gender, education, physical activity, body mass index. One study (Li et al., 2022) did not control for any measures of diabetes, and one study did not control for cardiovascular disease (Weinstein et al., 2023). Adhesion to a healthy diet was considered in three studies (Gonçalves et al., 2023; Bhave et al., 2024; Li et al., 2022). Across all studies other covariates included were income, deprivation, depression, alcohol consumption, family history of dementia, total energy intake, HbA1C, total cholesterol, blood pressure, atrialfibrillation and left ventricular hypertrophy. Table 1 displays the characteristics of the included studies.

### The Quality of Evidence and Risk of Bias in included studies

The NOS scores varied from six to eight across studies. One study was considered medium quality, with a higher risk of bias (Cardosa et al., 2022). The four other studies were rated as high quality, as NOS scores were either seven (n =1; Weinstein) or eight (n = 3; Li et al., 2022; Goncalves et al., 2023; Bhave et al., 2024). Please see supplementary tables 3 and 4 for NOS scoring details.

### Narrative synthesis

Three out of the five studies found a significant negative main effect of consuming UPF on the cognitive outcome of interest (Goncalves et al., 2023; Li et al., 2022; Bhave et al., 2024), whilst all studies highlighted a significant adverse consequence of consumption in either a sub-group of the population (Cardosa et al., 2022) or a sub-group of UPF type (Weinstein et al., 2023). Overall therefore, deleterious effects of UPF consumption on multiple cognitive health outcomes were identified across all studies but the combined results suggest that there may be specificity in the relationship between UPF and brain health in terms of participant characteristics (e.g. health status) and/or sub-type of UPF consumed. The main results of included studies are summarised in table 2.

**Table 2:** Analysis methods, main results and conclusions of included studies.

| Authors, Year | Outcome | Outcome measurement | Statistical analysis | Main Results | Conclusions |
| --- | --- | --- | --- | --- | --- |
| Cardoso et al., 2022 | Cognitive performance | Three cognitive tests: Consortium to Establish a Registry for Alzheimer's Disease (CERAD), Word Learning test, Animal Fluency test, and the Digit Symbol Substitution test (DSST) | Linear regression models | No significant association between UPF intake and performance on any cognitive test.<br>In those with no pre-existing diseases, individuals in the highest tertile of UPF consumption performed significantly worse on the Animal Fluency test ($\beta = -1.37$, 95% CI: -2.72; -0.03, $p = .046$) compared to the lowest consumption tertile. | UPF intake was not associated with cognitive performance in the full sample.<br>There may be an interaction between UPF consumption and health status as higher UPF consumption was associated with worse performance on the Animal Fluency task, but only among those with no CVD or diabetes. |
| Li et al., 2022 | All-cause dementia | Linkage with hospital admission data and death registers | Cox proportion -al hazards regression models | Increasing UPF intake by 10% was associated with 25% significant increase in the risk of all-cause dementia (HR: 1.25; 95% CI: 1.14–1.37; $p < .001$).<br>Replacing 10% of UPF weight in the diet with equivalent, but less processed foods, was estimated to be associated with a 19% lower risk of all-cause dementia (HR: 0.81; 95% CI 0.74–0.89; $p < 0.001$). | A higher UPF intake was significantly associated with a higher risk of all-cause dementia. Replacing UPF with less processed alternatives was associated with a reduced risk of developing dementia. |
| Weinstien et al., 2022 | Cognitive decline | 14 cognitive tests that were grouped into 4 domains. 1) Episodic memory: word list memory, word list recall, and word list recognition (CERAD). 2) Attention/working memory: the diamond cancelation and the digit span (forward and backward) tests from the Wechsler Memory Scale-Revised (WMS-R). 3) Semantic categorization: similarities, letter fluency and animal fluency tests. 4) Executive function: trail making test (A and B), CERAD-constructional praxis and digit symbol from the Wechsler Adult Intelligence Scale (WAIS)-Revised. Raw scores converted to Zscores . | Mixed-effect models with random intercepts and slopes | Total UPF consumption was not associated with cognitive change during the follow-up (of any domain).<br>Higher intake of UPF meat (Executive function: $\beta = -0.041$, SE: 0.013; $p = .002$ and Global Cognition: $\beta = -0.026$, SE: 0.010; $p = .011$) and oils/spreads was associated with greater decline in executive function and global cognition (oils and spreads Executive Function: $\beta = -0.037$, SE: 0.014; $p = .006$ and Global Cognition: $\beta = -0.028$, SE: 0.010; $p = .009$.) | In a type 2 diabetic cohort, total UPF intake was not associated with cognitive change.<br>Particular sub-groups of UPF may be harmful for cognitive health as ultra-processed meat and oils/spreads were associated with accelerated decline in executive functioning and global cognition. |
| Gonçalves et al., 2023 | Cognitive decline | 6 cognitive tests that were grouped into three domain composites. 1) Executive function: phonemic and semantic verbal fluency tests, and Trail-Making Test B. 2) | Linear mixed-effects models | Individuals with UPF intake above the lowest quartile of consumption exhibited a 28% ($\beta = -0.004$; 95% CI: -0.006 , -0.001; $P = .003$) faster rate of global cognitive | Higher consumption of UPFs was associated with a higher rate of global and executive function decline. Age and overall healthy |
| | | Memory: Immediate and delayed word recall, Word recognition (CERAD). 3) Global: all cognitive tests. Raw scores converted to Zscores. | with random intercepts and slopes | decline and a 25% ($\beta = -0.003$, 95% CI: $-0.005$, $0.000$; $P = .01$) faster rate of executive function decline compared to individuals in the lowest quartile of consumption. | eating patterns might modify the effect of UPF consumption on cognitive health. |
| Bhave et al., 2024 | Cognitive impairment & cognitive decline | Impairment was defined using performance relative to normative sample on memory (Word List Learning and Delayed Recall) and fluency assessments (Animal Fluency and Letter F Fluency). Incident cognitive impairment was defined as impairment on $\geq 1$ memory assessment and $\geq 1$ fluency assessment. | Cox proportion-al hazards models | A 10% increase in UPF intake was associated with a 16% higher risk of cognitive impairment (HR; 1.16; 95% CI: 1.09–1.24, $p < .001$).<br>Greater UPF intake was associated with increased cognitive decline in Word Learning ($\beta = -0.0006$; 95% CI, $-0.0009$ to $-0.0004$; $P < .0001$), Delayed Recall ($\beta = -0.0005$; 95% CI, $-0.0007$ to $-0.0003$; $P < .0001$), Animal Fluency ($\beta = -0.0005$; 95% CI, $-0.0007$ to $-0.0003$; $P < .0001$) and Letter Fluency ($\beta = -0.0004$; 95% CI, $-0.0007$ to $-0.0002$; $P < .0001$). | Higher UPF intake was associated with an increased risk of developing cognitive impairment.<br>Greater UPF consumption was associated with accelerated cognitive decline. |

One study (Li et al., 2022), assessed the relationship between UPF intake and the risk of developing dementia in 72,000 participants. They found that higher consumption of UPF was associated with a higher risk of dementia, Alzheimer’s Disease and vascular dementia. Specifically, a 10% increase in the percentage of UPFs in the diet was associated with a 25% increase in the risk of all-cause dementia, 14% risk of Alzheimer’s and 28% increase in the risk of developing vascular dementia. To explore the effect of replacing UPF items with a less processed alternative, the authors conducted substitution analysis which highlighted how replacing 10% of UPF intake with equivalent but less processed food, reduced the risk of dementia by 19% and vascular dementia by 22%. Similarly, higher UPF intake was linked to increased risk of adverse cognitive outcomes in the Bhave and colleagues (2024) study identifying that a 10% increase in UPF consumption was significantly associated with a 16% higher risk of developing cognitive impairment.

Another study examined the association between UPF consumption and cognitive performance cross-sectionally (Cardoso et al., 2022). In this study of 3632 US adults aged over 60 years old, no significant association was identified between UPF intake and performance on any of the three cognitive tests administered. However, in the participants without CVD or diabetes UPF intake was inversely associated with performance on the Animal Fluency Task (a measure of language and executive function).

The relationship between cognitive change and UPF intake was investigated by three studies (Bhave; Goncalves et al., 2023; Weinstein et al., 2023;). Two studies found a significant association between increasing UPF consumption and accelerated cognitive decline (Bhave et al., 2023; Goncalves et al., 2023) whilst one study (Weinstein et al., 2023) found no relationship between total UPF intake and cognitive change. However, Weinstein and colleagues (2023) did find a relationship between specific sub-groups of UPFs and cognitive change; higher intake of ultra-processed meat and oils/spreads was associated with significantly faster decline in executive functions and global cognition.

## Discussion

This systematic review provides an overview and evaluation of the evidence for the associations between UPF intake and cognitive health outcomes. The review included five studies which covered four different cognitive outcomes. The cognitive outcomes investigated were cognitive performance, cognitive decline, dementia risk and cognitive impairment risk. Whilst not all studies highlighted a significant main effect of UPF intake on outcome of interest, all studies identified adverse consequences of consumption in either a sub-group of the population or a sub-group of UPF type. However, conclusions should be drawn with caution due to the small number of studies available investigating UPF intake classified according to the NOVA framework and cognitive outcomes, and the range of cognitive outcomes investigated across the limited available studies.

Our results largely align with the two other systematic reviews in the field of UPF and cognitive health. In a recent meta-analysis of UPF items, high UPF intake was associated with an increased risk of dementia (Henney et al., 2024), and a systematic review concluded increased risk of Alzheimer’s Disease from increased consumption of UPF items (Claudino et al., 2024). These findings are based on reviews that included literature where total UPF intake, as defined by NOVA, was not explicitly measured.

Additionally, these reviews focused on only AD and dementia diagnosis. Nevertheless, the results are consistent with our overarching cognitive outcomes review, which specifically aggregated studies measuring total UPF intake using NOVA. Also, our review reflects previous review findings that have explored individual UPF items such as sugar sweetened beverages that concluded adverse cognitive health outcomes from consumption (Sun et al., 2022). Moreover, this current review echoes the broader literature of negative health consequences from increased UPF exposure (e.g. Lane et al., 2024).

Heterogeneity in results was observed amongst the studies investigating UPF consumption and cognitive change, with two out of three studies highlighting negative effects of UPF exposure on cognitive decline. The discrepancy in findings of Weinstein and colleagues (2023) analysis compared to the other studies may be attributable to the specific population included, which focused exclusively on individuals with type 2 diabetes. This explanation is supported by findings from Carodosa and colleagues (2022) as they highlight UPF intake was only associated with cognitive performance in individuals without pre-existing health conditions. This suggests the effect UPFs have on cognition may be modified by the overall health status of the participant, and therefore the lack of significant main effect in Weinstein and colleagues (2023) might be attributable to participants with type 2 diabetes. Further studies are required to explore the cognitive consequences of UPF intake in other and more diverse populations.

Three of the five included studies consider diet quality (i.e. healthy diet) in their investigations (Goncalves et al., 2023; Li et al., 2022; Bhave et al., 2024). Incorporating diet quality into analyses is imperative to aid the discussion of whether UPF intake confers independent adverse effects on health, or whether it is diet quality that mediates any relationship between UPFs and health outcomes. As UPFs often lack nutritional value and their intake is inversely associated with fruits, vegetables and legumes (Martini et al., 2021), it may simply be that UPFs are related to deleterious outcomes because they are associated with an overall unhealthy diet. The adverse effects of UPF exposure on cognitive outcomes remained whilst controlling for adherence to a healthy diet in two out of the three studies (Bhave et al., 2024; Li et al., 2022). The persistence of these associations implies UPFs independently impact cognitive outcomes and therefore it may be the nature and extent of processing that is having negative consequences. Opposingly, the negative effects of UPF intake on cognitive decline were isolated to only those who were consuming an unhealthy diet, with those who ate a healthy diet not experiencing the detrimental cognitive outcomes from UPF intake in another study (Goncalves et al., 2023), suggesting that diet quality modifies the association between UPF and brain health. Future studies could incorporate a measure of diet quality or pattern in the analysis to help explore whether the effect of consuming UPF on cognitive outcomes is independent - or not - from the quality of the diet.

The exact mechanisms by which UPF consumption might lead to negative health consequences, and specifically poor cognitive health, remains to be fully understood. Ultra-processed food intake may influence cognitive health via both direct and indirect mechanisms. Direct mechanisms include a lack of nutrients supplied by a high UPF diet (Martini et al., 2021) directly impacting brain structure and function (Gomez-Pinallia & Tyagi, 2013). Indirect mechanisms include how UPFs affect systematic health which can culminate in neuropathological changes. For instance, biological pathways implicated in brain health such as inflammation, oxidative stress and the gut microbiome are thought to be modulated by UPF exposure (Martinez et al., 2021). To illustrate, UPFs are linked to pro-inflammatory processes through modulating the composition of the gut microbiome (Zinocker & Lindset, 2018), and inflammation is thought of as a central pathological hallmark of both cognitive decline (Yaffe et al., 2003) and dementia (Heneka et al., 2015).

As evidence of the potential harmful effects of UPF consumption grows, researchers and public health organisations have been appealing for governmental action to help curb the rising intake. However, the small number of studies identified for this review highlights how additional research is required to investigate the cognitive consequences of UPF intake before being useable for the development of cognitive health guidelines and strategies. Additionally, this review – along with others in the broader literature of UPFs and health (e.g. Lane et al., 2024) – noted possible heterogeneity between UPF subgroups and health outcomes. Such disparity in associations highlight the need for nuances in the discussion of UPFs and raises the importance of additional research in order to limit any confusion caused by conflicted or altered public health campaigns.

Methodological differences between the five included studies should be noted. Firstly, four different cognitive outcomes were explored, as such the ability to collate information on the specific cognitive outcomes is limited. Second, different methods were employed to measure dietary intake across studies with some utilising 24-hour recalls and others using FFQs. Dietary intake methods are noteworthy due to the impact they have on the ability to classify food items as ultra-processed or not; the different tools collect varying specificity on food items eaten and how they are prepared and thus influence the precision of UPF classification. For example, 24-hr recalls allow the participant to freely record the intake of any food item along with the brand – this level of detail aids the assignment to the necessary NOVA category. Third, UPF intake was expressed as both a percentage of total calorie intake and as a percentage of total weight intake. Measuring relative UPF consumption in weight is suggested to more accurately capture UPF consumption compared to calorie intake due to the low-calorie nature of many UPFs (Rauber et al., 2018). However, the variation in exposure scores amongst included studies limits the capacity to compare across studies.

The strengths of this review primarily lie with the broad outcomes included coupled with the specificity in only NOVA classified UPF intake. As such, this review provides a targeted yet multifaceted overview of the current literature of UPF intake and all cognitive outcomes. Nonetheless, our review findings should be interpreted with caution due to several limitations. Principally, only five studies were included in the review due to the novelty of the research area; the limited number of studies therefore limits the generalisability, strength and confidence in the conclusions which can be drawn. Second, substantial heterogeneity existed between the studies. Variety was observed in terms of the cognitive outcome explored, the cognitive tests employed to measure cognitive function, the methods used to measure dietary intake (and thus exposure to UPF), the expression of UPF intake, along with other methodological differences. Third, as all included studies are of observational nature, this review does not attempt to determine causality but we invite future carefully designed RCTs.

## Conclusion

Adverse associations between UPF consumption and cognitive outcome were identified in all studies, if for some studies these negative associations were isolated to a sub-group of the population or a sub-group of UPF type. Due to the novelty of this research area, more studies are required to help elucidate whether, and how, UPF may affect cognitive health. Additionally, future analyses should incorporate a measure of overall diet quality to aid in determining whether the effect of UPF is independent of dietary pattern or influenced by it.

## Data Availability

All data produced in the present work are contained in the manuscript.

## References

Baker, P., Machado, P., Santos, T., Sievert, K., Backholer, K., Hadjikakou, M., … & Lawrence, M. (2020). Ultra-processed foods and the nutrition transition: Global, regional and national trends, food systems transformations and political economy drivers. Obesity Reviews, 21(12), e13126.

Bhave, V. M., Oladele, C. R., Ament, Z., Kijpaisalratana, N., Jones, A. C., Couch, C. A., … & Kimberly, W. T. (2024). Associations Between Ultra-Processed Food Consumption and Adverse Brain Health Outcomes. Neurology, 102(11), e209432.

Cardoso, B., Machado, P., & Steele, E. M. (2022). Association between ultra-processed food consumption and cognitive performance in US older adults: a cross-sectional analysis of the NHANES 2011-2014. European journal of nutrition, 61(8), 3975–3985. 10.1007/s00394-022-02911-1

Chang, K., Gunter, M. J., Rauber, F., Levy, R. B., Huybrechts, I., Kliemann, N., … & Vamos, E. P. (2023).

Cuevas-Sierra, A., Milagro, F. I., Aranaz, P., Martínez, J. A., & Riezu-Boj, J. I. (2021). Gut microbiota differences according to ultra-processed food consumption in a Spanish population. Nutrients, 13(8), 2710.

Claudino, P. A., Bueno, N. B., Piloneto, S., Halaiko, D., Azevedo de Sousa, L.P., Barroso Jara Maia, C.H., & Netto, B. D. M. (2024). Consumption of ultra-processed foods and risk for Alzheimer’s disease: a systematic review. Frontiers in nutrition, 10, 1288749. 10.3389/fnut.2023.1288749

Cuevas-Sierra, A., Milagro, F. I., Aranaz, P., Martínez, J. A., & Riezu-Boj, J. I. (2021). Gut Microbiota Differences According to Ultra-Processed Food Consumption in a Spanish Population. Nutrients, 13(8), 2710. 10.3390/nu13082710

Delpino, F. M., Figueiredo, L. M., Bielemann, R. M., da Silva, B. G. C., Dos Santos, F.S., Mintem, G. C., Flores, T. R., Arcêncio, R. A., & Nunes, B. P. (2022). Ultra-processed food and risk of type 2 diabetes: a systematic review and meta-analysis of longitudinal studies. International journal of epidemiology, 51(4), 1120–1141. 10.1093/ije/dyab247

Dicken, S. J., Dahm, C. C., Ibsen, D. B., Olsen, A., Tjønneland, A., Louati-Hajji, M., … & Batterham, R. L. (2024). Food consumption by degree of food processing and risk of type 2 diabetes mellitus: a prospective cohort analysis of the European Prospective Investigation into Cancer and Nutrition (EPIC). The Lancet Regional Health–Europe, 46.

GBD 2019 Dementia Forecasting Collaborators (2022). Estimation of the global prevalence of dementia in 2019 and forecasted prevalence in 2050: an analysis for the Global Burden of Disease Study 2019. The Lancet. Public health, 7(2), e105–e125. 10.1016/S2468-2667(21)00249-8

Gomez-Pinilla F. and Tyagi E., Diet and cognition: interplay between cell metabolism and neuronal plasticity, Current Opinion in Clinical Nutrition and Metabolic Care. (2013) 16, no. 6, 726–733.

Gonçalves, N. G., Ferreira, N. V., Khandpur, N., Steele, E. M., Levy, R. B., Lotufo, P. A., … & Suemoto, C. K. (2023). Association between consumption of ultraprocessed foods and cognitive decline. JAMA neurology, 80(2), 142–150.

Heneka, M. T., Carson, M. J., El Khoury, J., Landreth, G. E., Brosseron, F., Feinstein, D. L., … & Kummer, M. P. (2015). Neuroinflammation in Alzheimer’s disease. The Lancet Neurology, 14(4), 388–405.

Hu, F. B. (2011). Globalization of diabetes: the role of diet, lifestyle, and genes. Diabetes care, 34(6), 1249–1257.

Henney, A. E., Gillespie, C. S., Alam, U., Hydes, T. J., Mackay, C. E., & Cuthbertson, D. J. (2024). High intake of ultra-processed food is associated with dementia in adults: a systematic review and meta-analysis of observational studies. Journal of neurology, 271(1), 198–210. 10.1007/s00415-023-12033-1

Lane, M. M., Gamage, E., Du, S., Ashtree, D. N., McGuinness, A. J., Gauci, S., … & Marx, W. (2024). Ultra-processed food exposure and adverse health outcomes: umbrella review of epidemiological meta-analyses. bmj, 384.

Li, H., Li, S., Yang, H., Zhang, Y., Zhang, S., Ma, Y., Hou, Y., Zhang, X., Niu, K., Borné, Y., & Wang, Y. (2022). Association of Ultraprocessed Food Consumption With Risk of Dementia: A Prospective Cohort Study. Neurology, 99(10), e1056–e1066. 10.1212/WNL.0000000000200871

Livingston, G., Huntley, J., Liu, K. Y., Costafreda, S. G., Selbæk, G., Alladi, S., Ames, D., Banerjee, S., Burns, A., Brayne, C., Fox, N. C., Ferri, C. P., Gitlin, L. N., Howard, R., Kales, H. C., Kivimäki, M., Larson, E. B., Nakasujja, N., Rockwood, K., Samus, Q., … Mukadam, N. (2024). Dementia prevention, intervention, and care: 2024 report of the Lancet standing Commission. Lancet (London, England), 404(10452), 572–628. 10.1016/S0140-6736(24)01296-0

Martínez Leo, E.E., Peñafiel, A. M., Hernández Escalante, V.M., & Cabrera Araujo, Z. M. (2021). Ultra-processed diet, systemic oxidative stress, and breach of immunologic tolerance. Nutrition (Burbank, Los Angeles County, Calif.), 91-92, 111419. 10.1016/j.nut.2021.111419

Martini, D., Godos, J., Bonaccio, M., Vitaglione, P., & Grosso, G. (2021). Ultra-processed foods and nutritional dietary profile: a meta-analysis of nationally representative samples. Nutrients, 13(10), 3390.

Monteiro, C. A., Cannon, G., Moubarac, J. C., Levy, R. B., Louzada, M. L. C., & Jaime, P. C. (2018). The UN Decade of Nutrition, the NOVA food classification and the trouble with ultra-processing. Public health nutrition, 21(1), 5–17.

Nichols, E., Steinmetz, J. D., Vollset, S. E., Fukutaki, K., Chalek, J., Abd-Allah, F., … & Liu, X. (2022). Estimation of the global prevalence of dementia in 2019 and forecasted prevalence in 2050: an analysis for the Global Burden of Disease Study 2019. The Lancet Public Health, 7(2), e105–e125.

Rauber, F., da Costa Louzada, M.L., Steele, E. M., Millett, C., Monteiro, C. A., & Levy, R. B. (2018). Ultra-Processed Food Consumption and Chronic Non-Communicable Diseases-Related Dietary Nutrient Profile in the UK (2008?2014). Nutrients, 10(5), 587. 10.3390/nu10050587

Rauber, F., Chang, K., Vamos, E. P., da Costa Louzada, M.L., Monteiro, C. A., Millett, C., & Levy, R. B. (2021). Ultra-processed food consumption and risk of obesity: a prospective cohort study of UK Biobank. European journal of nutrition, 60(4), 2169–2180. 10.1007/s00394-020-02367-1.

Rico-Campà, A., Martínez-González, M. A., Alvarez-Alvarez, I., de Deus Mendonça, R., de la Fuente-Arrillaga, C., Gómez-Donoso, C., & Bes-Rastrollo, M. (2019). Association between consumption of ultra-processed foods and all cause mortality: SUN prospective cohort study. bmj, 365.

Silva Dos Santos, F., Costa Mintem, G., Oliveira de Oliveira, I., Lessa Horta, B., Ramos, E., Lopes, C., & Petrucci Gigante, D. (2023). Consumption of ultra-processed foods and IL-6 in two cohorts from high- and middle-income countries. The British journal of nutrition, 129(9), 1552–1562. 10.1017/S0007114522000551

Srour, B., Fezeu, L. K., Kesse-Guyot, E., Allès, B., Méjean, C., Andrianasolo, R. M., … & Touvier, M. (2019). Ultra-processed food intake and risk of cardiovascular disease: prospective cohort study (NutriNet-Santé). bmj, 365.

Sun, Q., Yang, Y., Wang, X., Yang, R., & Li, X. (2022). The association between sugar-sweetened beverages and cognitive function in middle-aged and older people: A meta-analysis. The Journal of Prevention of Alzheimer’s Disease, 9(2), 323–330.

Weinstein, G., Vered, S., Ivancovsky-Wajcman, D., Ravona-Springer, R., Heymann, A., Zelber-Sagi, S., Shahar, D. R., & Beeri, M. S. (2023). Consumption of Ultra-Processed Food and Cognitive Decline among Older Adults With Type-2 Diabetes. The journals of gerontology. Series A, Biological sciences and medical sciences, 78(1), 134–142.

Wells, G. A., Shea, B., O’Connell, D., Peterson, J., Welch, V., Losos, M., & Tugwell, P (2015). The Newcastle-Ottawa Scale (NOS) for assessing the quality of nonrandomised studies in meta-analyses.

World Health Organization. (2023). Dementia. https://www.who.int/news-room/fact-sheets/detail/dementia

Yaffe K, Lindquist K, Penninx BW. et al. Inflammatory markers and cognition in well-functioning African-American and white elders. Neurology. 2003;61:76–8012847160

Zinöcker, M. K., & Lindseth, I. A. (2018). The Western diet–microbiome-host interaction and its role in metabolic disease. Nutrients, 10(3), 365.

Zhang, Z., Jackson, S. L., Martinez, E., Gillespie, C., & Yang, Q. (2021). Association between ultraprocessed food intake and cardiovascular health in US adults: a cross-sectional analysis of the NHANES 2011–2016. The American journal of clinical nutrition, 113(2), 428–436.

