## Supplemental files for "Ultra-processed food exposure and cognitive outcomes: A systematic review of observational studies"

**Supplementary Materials**

Supplementary table 1: Inclusion and exclusion criteria used in the selection of papers identified in the systematic search, to be included in this review.

Inclusion criteria:

| Item | Definition |
| --- | --- |
| Study design | Original research studies published in a peer reviewed journal. Studies to be of observational design (cross-sectional, prospective cohort, case-control). |
| Exposure | Had to assess UPF intake, defined using the NOVA food classification system (must make a direct reference to UPFs and NOVA).  The study must measure overall dietary intake of UPFs (not measure the intake of a single UPF item). |
| Outcome | Must assess the association of UPF with a cognitive outcome (dementia, (mild) cognitive impairment, cognitive performance (of any cognitive domain), cognitive performance trajectories).  Dementia includes AD, VD, frontotemporal dementia or Lewy Body dementia (does not include early onset dementia). The definition of dementia had to be based on diagnostic criteria or data/hospital linkage. |
| Effect Size | Odds Ratio; Relative Risk; Hazards Ratio / beta coefficients (when assessing cognitive performance). |
| Participants | Human adult participants (over 18 years old) |

Exclusion criteria:

| Item | Definition |
| --- | --- |
| Study design | Animal studies. Reviews, case-reports, case series, conference abstracts, book chapters, editorials or letters, non- English language. |
| Exposure | Studies which measured the intake of only one type of UPFs. Studies that determine the processing level of foods using any other method than the NOVA classification system. Studies which assessed dietary patterns which (post hoc) are high in UPFs but did not directly assessing the relationship between UPFs and (cognitive) outcome. |
| Outcome | Non-dementia neurological disorders (e.g. stroke, Parkinsons, epilepsy). |
| Participants | Human participants under 18 years old. Non-human participants. |

Supplementary Table 2 : Full search strategies by database.

| Database | Search terms | Number articles returned |
| --- | --- | --- |
| MEDLINE, APA Psych Articles, PsycINFO and Embase through OVID | #1. ultra-processed food.ab. or ultra-processed food.ti. or ultraprocessed food.ab. or ultraprocessed food.ti. or ultra processed food.ab. or ultra processed food.ti. or UPF.ab. or UPF.ti. or NOVA.ab. or NOVA.ti.    #2. Dementia.ab. or Dementia.ti. or Alzheimer.ab. or Alzheimer.ti. or Lewy body.ab. or Lewy body.ti. or LBD.ab. or LBD.ti. or frontotemporal.ab. or frontotemporal.ti. or cognitive.ab. or cognitive.ti. or cognition.ab. or cognition.ti. or MCI.ab. or MCI.ti. or memory.ab. or memory.ti. or brain function.ab. or brain function.ti. or executive function.ab. or executive function.ti.    #3. 1 AND 2 | 22157          2918863        509 |
| PUBMED | #1. "Dementia"[Title/Abstract] OR "alzheimer"[Title/Abstract] OR "lewy body"[Title/Abstract] OR "LBD"[Title/Abstract] OR "frontotemporal"[Title/Abstract] OR "cognitive"[Title/Abstract] OR "cognition"[Title/Abstract] OR "MCI"[Title/Abstract] OR "memory"[Title/Abstract] OR "brain function"[Title/Abstract] OR "executive function"[Title/Abstract]    #2. "ultra processed food"[Title/Abstract] OR "ultra-processed food"[Title/Abstract] OR "ultraprocessed food"[Title/Abstract] OR "UPF"[Title/Abstract] OR "NOVA"[Title/Abstract]    3#. 1 AND 2 | 1,024,487          8,717      129 |

Supplementary Table 3: Quality of evidence of included studies reported via the Newcastle Ottawa Quality Assessment Scale for longitudinal studies.

| **Author, year** | Li et al., 2022 | Weinstien et al., 2023 | Gonçalves et al., 2023 | Bhave et al., 2024 |
| --- | --- | --- | --- | --- |
| **NOS component** |  |  |  |  |
| Representativeness of exposed cohort | a | a | a | a |
| Selection of non-exposed cohort | a | a | a | a |
| Ascertainment of exposure | c | c | c | c |
| Outcome of interest not present at start of study | a | a | a | a |
| Comparability of cohorts | a & b | a & b | a & b | a & b |
| Assessment of outcome | a | a | a | a |
| Follow up length | a | a | a | a |
| Follow up rate | b | b | b | c |
| **Total Score** | **8** | **8** | **8** | **7** |

Supplementary Table 4: Quality of evidence of included studies reported via the Newcastle Ottawa Quality Assessment Scale for cross-sectional studies.

| **Authors, Year** | **Representativeness of sample** | **Sample size** | **Non-respondents** | **Ascertainment of exposure** | **Confounding factors controlled.** | **Ascertainment of outcome** | **Statistics** | **Total score** |
| --- | --- | --- | --- | --- | --- | --- | --- | --- |
| Cardoso et al., 2022 | 1 | 0 | 0 | 0 | 2 | 2 | 1 | **6** |
